# *Stenotrophomonas muris* - First discovered as an urgent human pathogen with strong virulence associated with bloodstream infections

**DOI:** 10.1101/2024.05.16.24307319

**Authors:** Jiaying Liu, Xu Dong, Yuyun Yu, Tiantian Wu, Yanghui Xiang, Xin Yuan, Dan Cao, Kefan Bi, Hanyin Zhang, Lixia Zhu, Ying Zhang

## Abstract

For the first time, *Stenotrophomonas muris* (*S. muris*) has been identified to be associated with human infections while studying the virulence of *Stenotrophomonas maltophilia* (*S. maltophilia*) clinical isolates. Previously, *S. muris* was only isolated from the intestines of mice but its pathogenic potential for humans has never been reported. In this work, the phenotype of *S. muris* virulence, the potential genes that encode higher virulence of *S. muris*, and host responses to *S. muris* infection were investigated for the first time. It was found that S9 (*S. muris* no.9, isolated from patient’s bloodstream infection) was more virulent than both S8 (*S. muris* no.8, isolated from patient’s sputum) and S1 (*S. maltophilia*), where S8 and S9 were subsequently identified as *S. muris* by whole genome sequencing analysis. Candidate genes which may encode higher virulence of S9 were identified, including *virB6, dcm, hlyD*, and 14 other genes involved in porphyrin metabolism, pyrimidine metabolism, DNA methylation, two component system, and biofilm formation. Transcriptome analysis of host cells (THP-1 cells) infected by S8 and S9 (S9 with strong virulence over S8 with weak virulence) showed that 12 candidate genes involved in ion transport function and calcium signaling pathway were down-regulated and require special attention. Antibiotic susceptibility testing indicated that compared with *S. maltophilia*, the *S. muris* strains, though more susceptible to minocycline, are highly resistant to last resort antibitoics colistin and polymyxin B and are also resistant to cephalosporin and fluoroquinolone. Because of the above differences in virulence properties and antibiotic susceptibility, it is critical that *S. muris* be distinguished from *S. maltophilia* in clinical setting for improved care. This work provides the basis for future studies on pathogenic mechanisms of *S. muris* and for developing improved treatment in the future.

## Introduction

According to the World Health Organization (WHO), infectious diseases rank fourth among the top ten causes of death globally [1]. It is well known that pathogenic bacteria are one of the main culprits of infectious diseases. It is estimated that thousands of bacterial species have been discovered on Earth, but only a few hundred can cause human disease. The key difference between the bacteria that can cause disease and those that cannot is that the former can produce virulence factors that harm humans. Virulence factors help bacteria enter the host and form a beneficial niche for the invading bacteria. They can also deactivate the host defence system so the immune response becomes weakened. They also help bacteria to multiply in the host and to spread to other hosts. Although bacterial virulence is either strong or weak, we should not only care about the strong ones but also pay attention to the weak ones; since even if a bacterium has weak virulence, it may be strongly harmful to humans under immunocompromised conditions as in opportunistic pathogens. Opportunistic pathogens are normal bacteria in the host that can usually maintain a good survival balance, but cause infections when the balance is broken. An example of the opportunistic pathogen is *Stenotrophomonas maltophilia* (*S. maltophilia*), which is of weak virulence but may cause high mortality rate. Therefore, when studying a newly discovered pathogen, characterizing its virulence should be one of the primary research objectives.

Unfortunately, bacterial virulence is usually complex because of its high context-dependence. The antibiotic levels, pathogen types and other factors such as the pH and oxidative stress in the host may have impacts on bacterial virulence. For example, in pulmonary infections, *P. aeruginosa* and COVID-19 may enhance the virulence of *S. maltophilia* and show cooperative pathogenicity [2]. Biofilm formation can affect bacterial virulence. It has been reported that synergistic multispecies biofilms were formed when several opportunistic pathogens were growing in the same environment [3]. The complexity of the bacterial virulence brings significant challenges for study. The good news is that bacteria virulence is usually encoded in the genes, so the fast-developing genetic and genomics tools can provide great aid in study of bacterial virulence. With the help of bioinformatics and mutant library screens, and comparative genome sequencing of bacterial pathogens and their non-pathogenic counterparts can help identify the virulence factors of a pathogenic bacterium [4].

In this work, we discovered that two clinical isolates named S8 and S9, which were initially identified as *S. maltophilia* by standard lab microbial identification by mass spectrometry, had stronger virulence than the *S. maltophilia* type strain ATCC13637 (named S1). S8 was isolated from a patient sputum sample and S9 was from the bloodstream infection of another patient. The two more virulent isolates were subsequently recognized as *Stenotrophomonas muris* (*S. muris*) [5] by whole genome sequencing (WGS) analysis. Here we report the virulence properties and whole genome sequence analysis of *S. muris* strains in comparison with *S. maltophilia* as well as unique features of host response to *S. muris* infection.

## Results and Discussion

### Mass spectrometry and biochemical testing misidentified two S. muris clinical isolates as S. maltophilia

In routine clinical microbiology identification of clinical isolates, matrix assisted laser desorption/ionization-time of flight mass spectrometry (MALDI-TOF MS) (Vitek MS system, bioMerieux, France), identified strains S8 and S9 as *S. maltophilia*. Then, identification by biochemical tests using bioMerieux API 20E kit showed the same results (Table 1). According to the database, the probability that S8 and S9 are *S. maltophilia* is 99.3%.

**Table 1.**
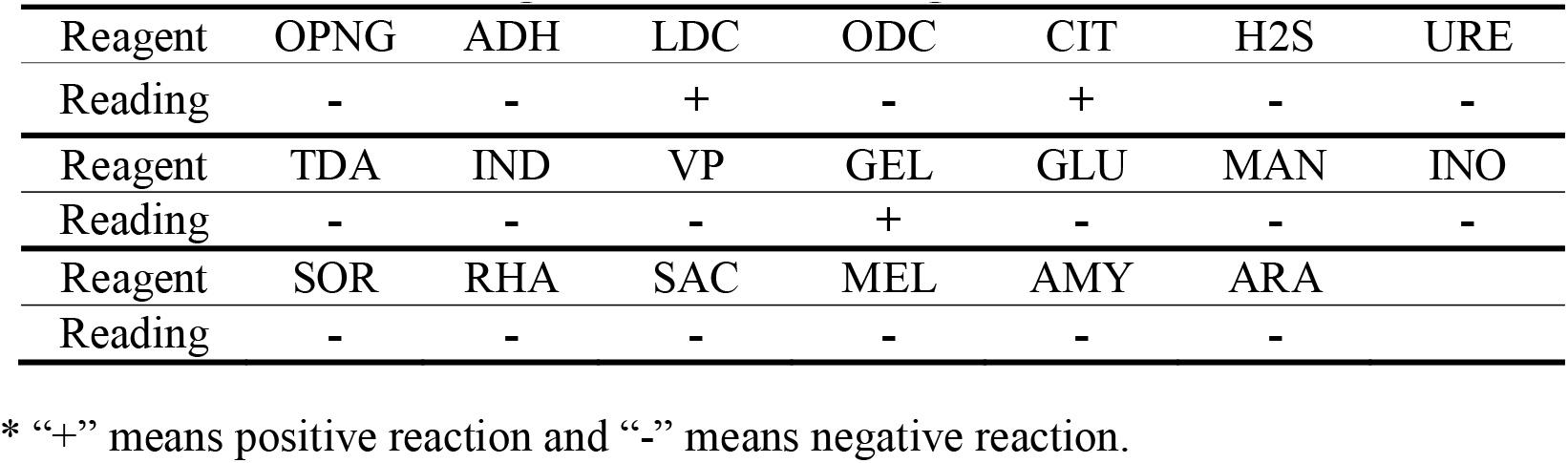
Biochemical testing results of the 20 reagents for strains S8 and S9*.

### SYTOX Green staining and Lactate dehydrogenase (LDH) release assay

To determine the cytotoxicityof the isolated strains, the human monocytic leukaemia (THP-1) cells were infected with S8, S9 and S1 for 8 hours (h) (Fig. 1a) and 18h (Fig. 1b), respectively followed by staining with SYTOX Green. In both Figs. 1a and 1b, the control groups (CT) had lowest staining level as expected. And the ranks of the staining levels for both Figs. 1a and 1b are S9>S8>S1>CT, which implies the rank of the virulence is S9>S8>S1. The comparison between Fig. 1a and Fig. 1b also implies that longer bacterial infections resulted in higher mortality of the THP-1 cells, especially for cells infected with S9.

**Figure 1.**
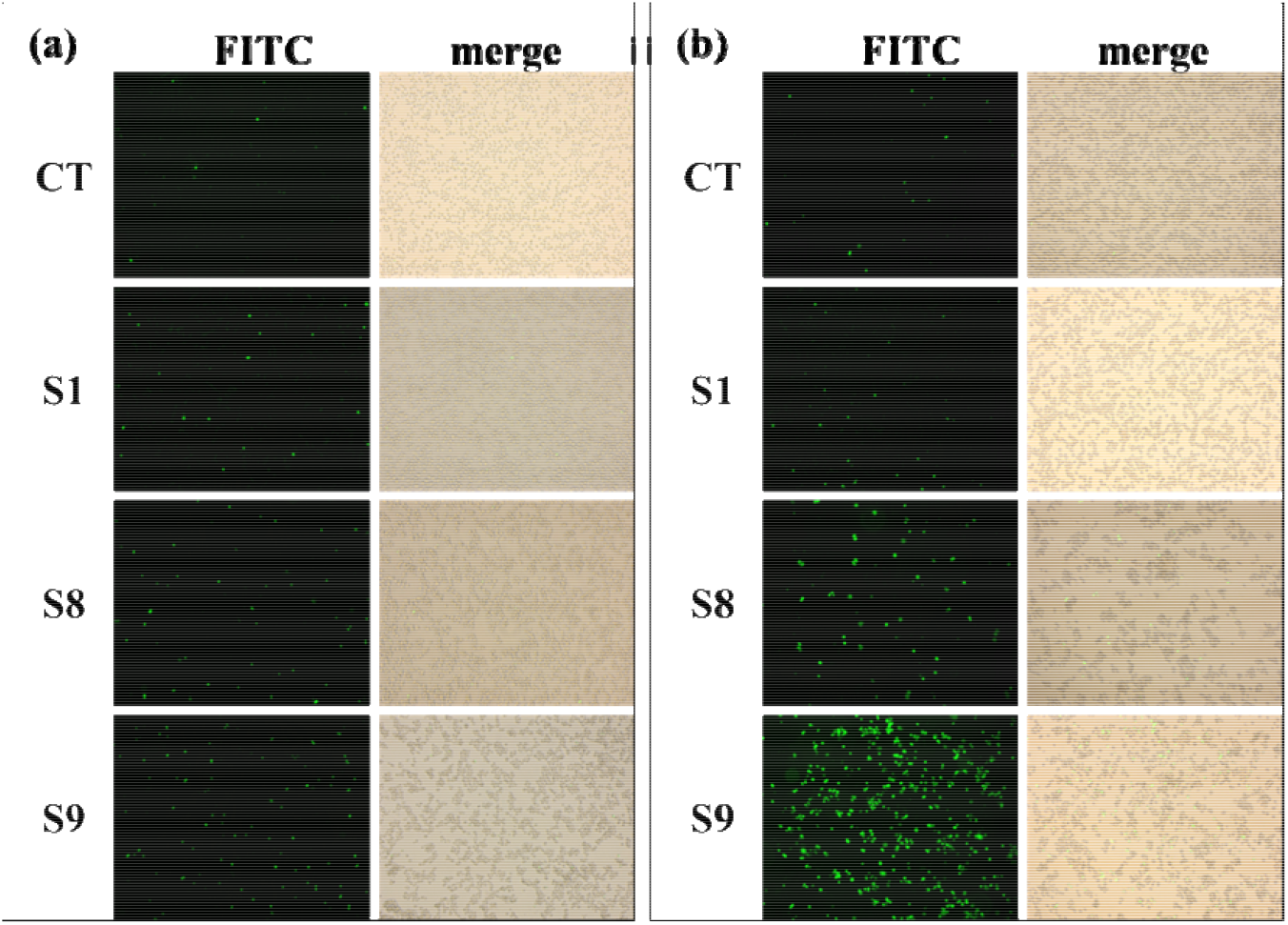
SYTOX Green staining of THP-1 infected cells. THP-1 cells were infected by S1, S8, or S9 bacteria for (a) 8h and (b) 18h, and observed under a 10× microscope. “FITC” were shot in fluorescence mode and “merge” were merged pictures of “FITC” and those shot in normal mode. From the “merge” pictures, the proportion of dead cells could be read. S1, S8 and S9 in the figure were THP-1 cells infected by S1, S8 and S9, respectively. CT was control THP-1 cells without infection.

LDH release assay to reflect the degree of cell death upon infection of the THP-1 cells by bacterial strains can also be used to quantitate bacterial virulence. As shown in Fig. 2, the death rates of the THP-1 cells infected by S1, S8 and S9 were 3.24%±0.88%, 14.08%±5.26% and 22.93%±7.39%, respectively. The mortality caused by S9 was about 7 times of that caused by S1, which means S9 is much more virulent than S1. And the virulence rank would be S9>S8>S1, just the same as that obtained from the above SYTOX Green staining.

**Figure 2.**
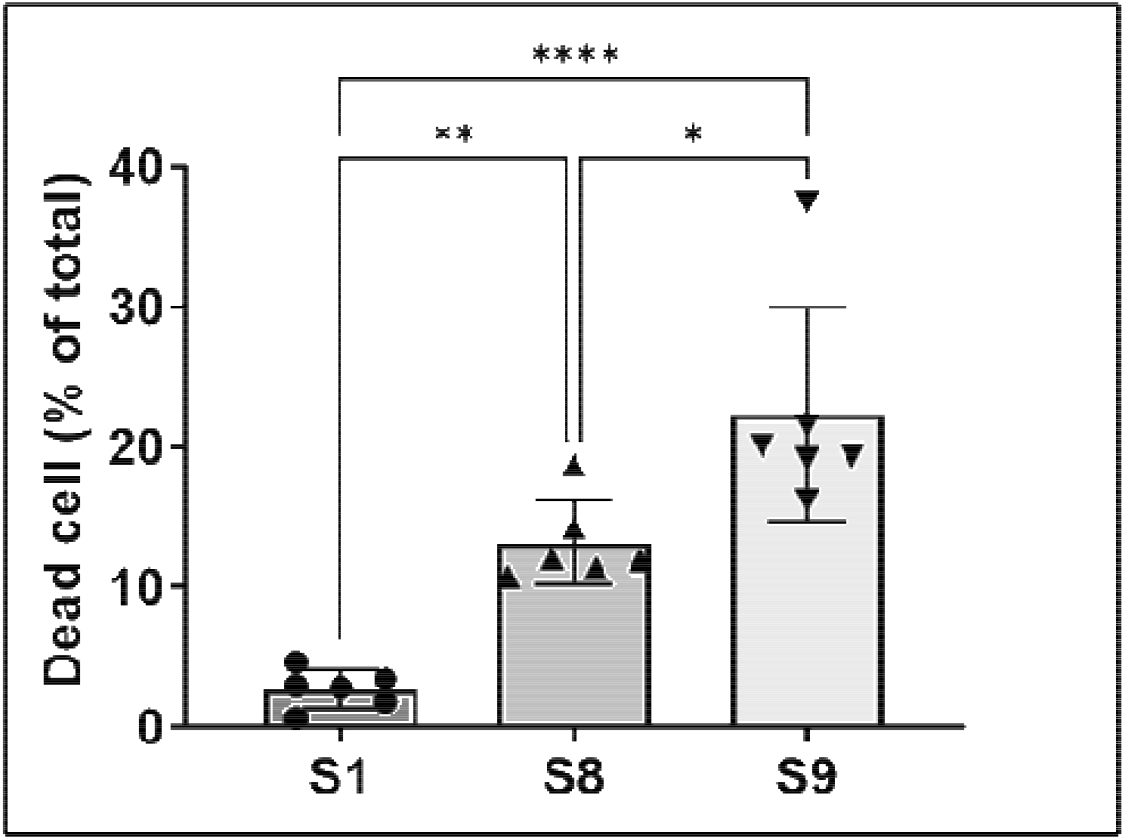
LDH release of infected THP-1 cells. LDH release of THP-1 cells after 24h infection with S1, S8, or S9 at MOI=1:1. The death rates of the infected cells were calculated according to M_cell_ (%) = (A_t_ - A_c_) / (A_e_ - A_c_) × 100%, where M_cell_ was the death rate, A_t_ was the absorbance (at 490nm) of the cells infected by the bacteria, A_c_ was the absorbance of the culture suspension, A_e_ was the absorbance of the all-lytic cells (maximum absorbance). S1, S8 and S9 were group names of THP-1 cells infected by S1, S8 and S9, respectively.

### Survival curves of infected G. mellonella larvae

Survival curves of infected *G. mellonella* larvae can quantitate in vivo bacterial virulence. In Fig. 3a, the gross view of the health status of the *G. mellonella* larvae infected by no infection, PBS, or S1, S8, and S9 after 7 days is presented. As can be seen, they had no death for uninfected or injected with PBS, and 2, 4, 7 dead larvae in S1, S8, and S9 infected groups, respectively, resulting in rank of virulence S9>S8>S1. In Fig. 3b, the 7-day survival curves of five groups (10 in each group) of *G. mellonella* larvae were plotted. The two control groups were *G. mellonella* larvae infected by no bacteria and by PBS. Fig. 3(b) also shows that *G. mellonella* larvae infected by S9 died more rapidly than that infected by S1, and the 7-day survival rates of the *G. mellonella* larvae infected by S1, S8 and S9 were 80%, 60% and 30%, respectively, which implies the virulence rank is also S9>S8>S1.

**Figure 3.**
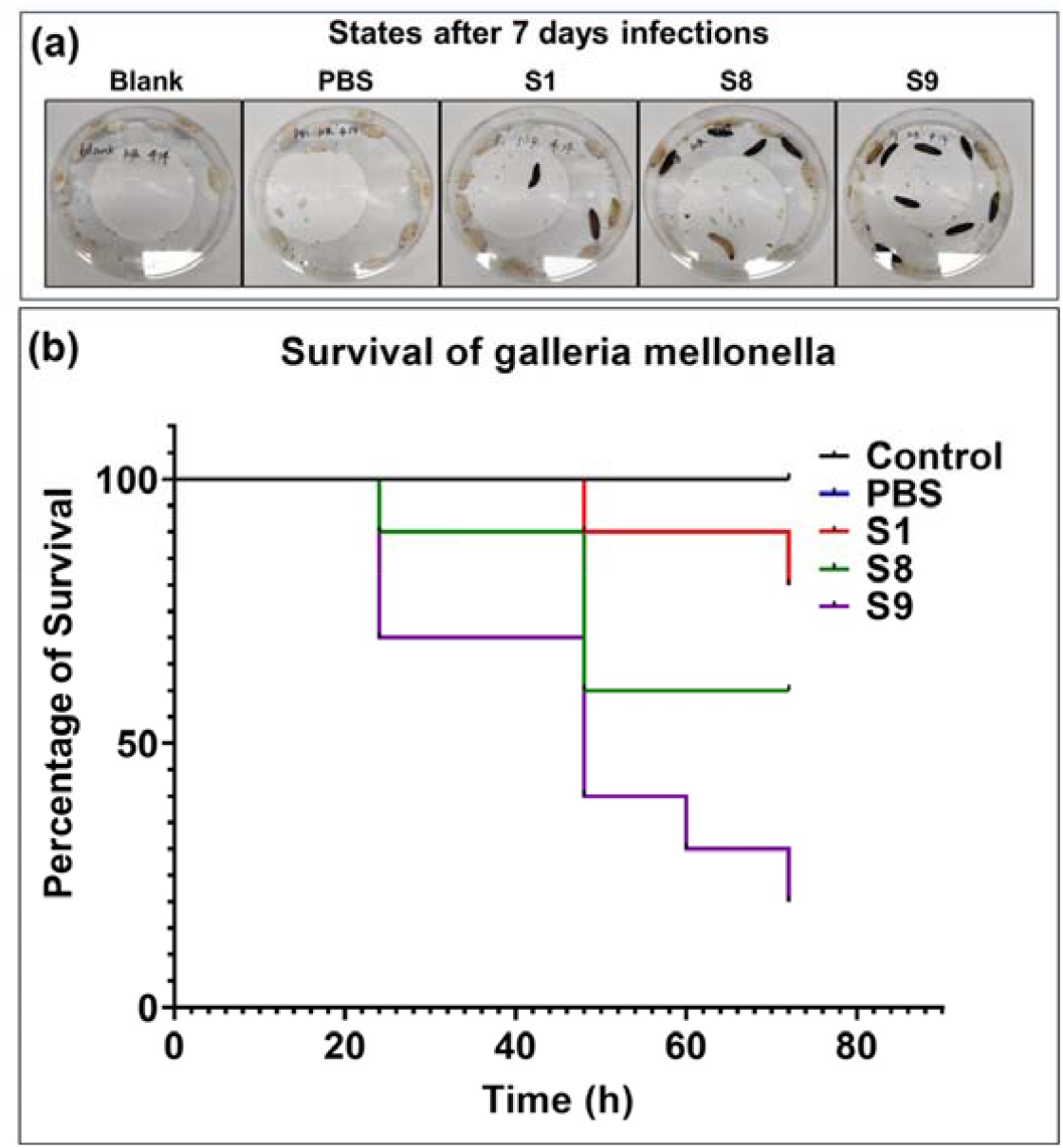
Survival curves of *G. mellonella* larvae infected with different bacterial strains or control. In the control group, the larvae were infected by nothing. In the PBS group, the larvae were injected with PBS buffer. In S1, S8 and S9 groups, the larvae (10 per group) were infected by S1, S8 and S9, respectively. The ordinate is percentage of survival.

### Survival curves of mice infected with S. muris and S. maltophilia strains

Survival curves for mice infected with the *S. muris* and *S. maltophilia* strains were also employed because mouse is a more relevant mammalian model. The survival curves of three groups (10 in each group) of mice (infected by S1, S8 and S9) are shown in Fig. 4. As can be seen in Fig. 4, S9 exhibited a rather strong virulence because all mice in this group rapidly died within 2 days. The survival rates of the mice infected by S1, S8, S9 were 10%, 60% and 0%, respectively, again indicating that S9 is the most virulent.

**Figure 4.**
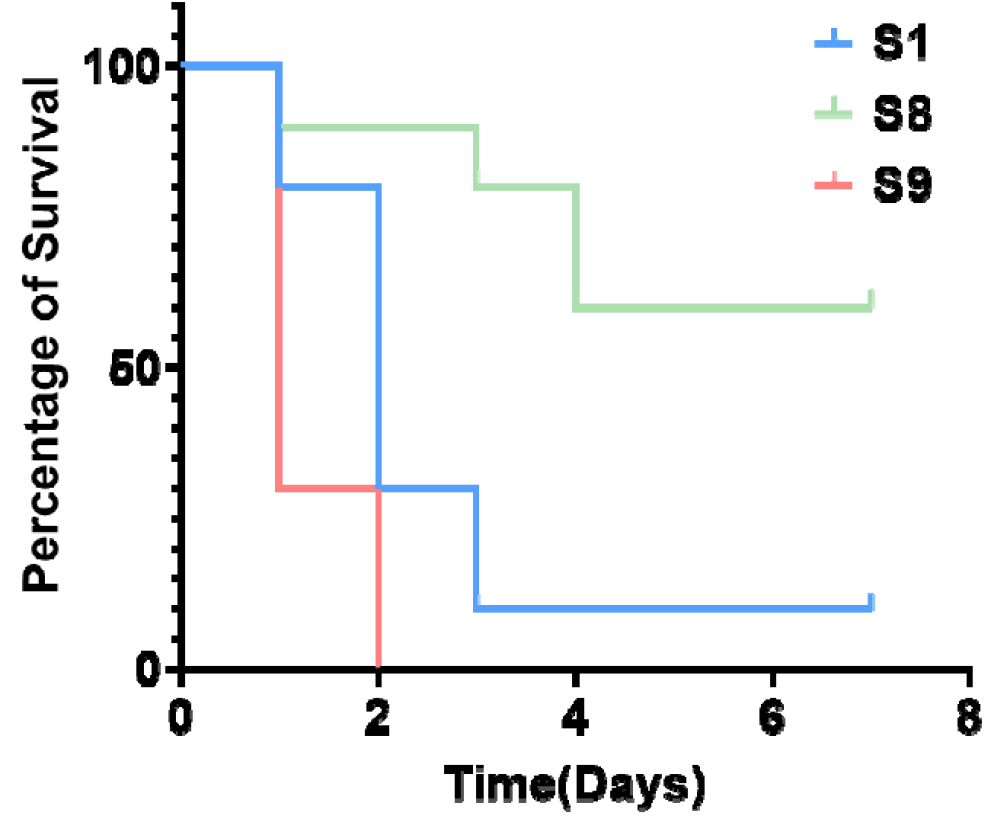
Survival of infected mice. In S1, S8 and S9 groups, the mice (10 per group) were infected by S1, S8 and S9 (1×10^8^ CFU per mouse), respectively and observed for survival or death over 7 days. The ordinate is percentage of survival.

### WGS and average nucleotide identity (ANI)

WGS showed that the S9 genome is composed of 4,608,528 bp with the Guanine-Cytosine (GC) content 66.8%. The whole genome of S9 contains a chromosome with 4,507,625 bp (66.8% GC) and a plasmid with 91,348 bp (67.7% GC). By analyzing the WGS data via ANI method, it was shown that S8 and S9 were ∼99% consistent with *S. muris*, but only ∼92% consistent with *S. maltophilia*, which means both S8 and S9 were *S. muris*. Deeper analysis showed that there were 4280 genes in S8 and 4271 genes in S9 (4197 genes in chromosome and 74 genes in plasmid).

### In silico analysis of vlirulence factors of S9 in the Virulence Factor Database (VFDB) and pathway enrichment analyses

All S9 genes were compared with the VFDB and 33 S9 genes (with the identity >60%) were found in VFDB. The 33 genes are listed in Table S1 in Supplementary Materials. In Table S1, S8 genes and S1 genes annotated in VFDB are also listed as the unique genes of S9 are regarded the candidate genes that may encode higher virulence of S9. S9 genes annotated in VFDB repeated by S1 or S8 are excluded as the candidate genes in Table S1.

We also performed gene enrichment analysis with gene ontology (GO) database and Kyoto Encyclopaedia of Genes and Genomes (KEGG) database. The target genes for enrichment were S9-unique genes, which were genes in S9 but not in S8. Since S1 is also less virulent than S9, these S9-unique genes versus S1 were excluded. There were 280 S9-unique genes, and 19 of them were in S1. And in the remaining 261 S9-unique genes, 53 (listed in Table S2) were annotated in GO database and 14 (Table 2) were annotated in KEGG database.

**Table 2.**
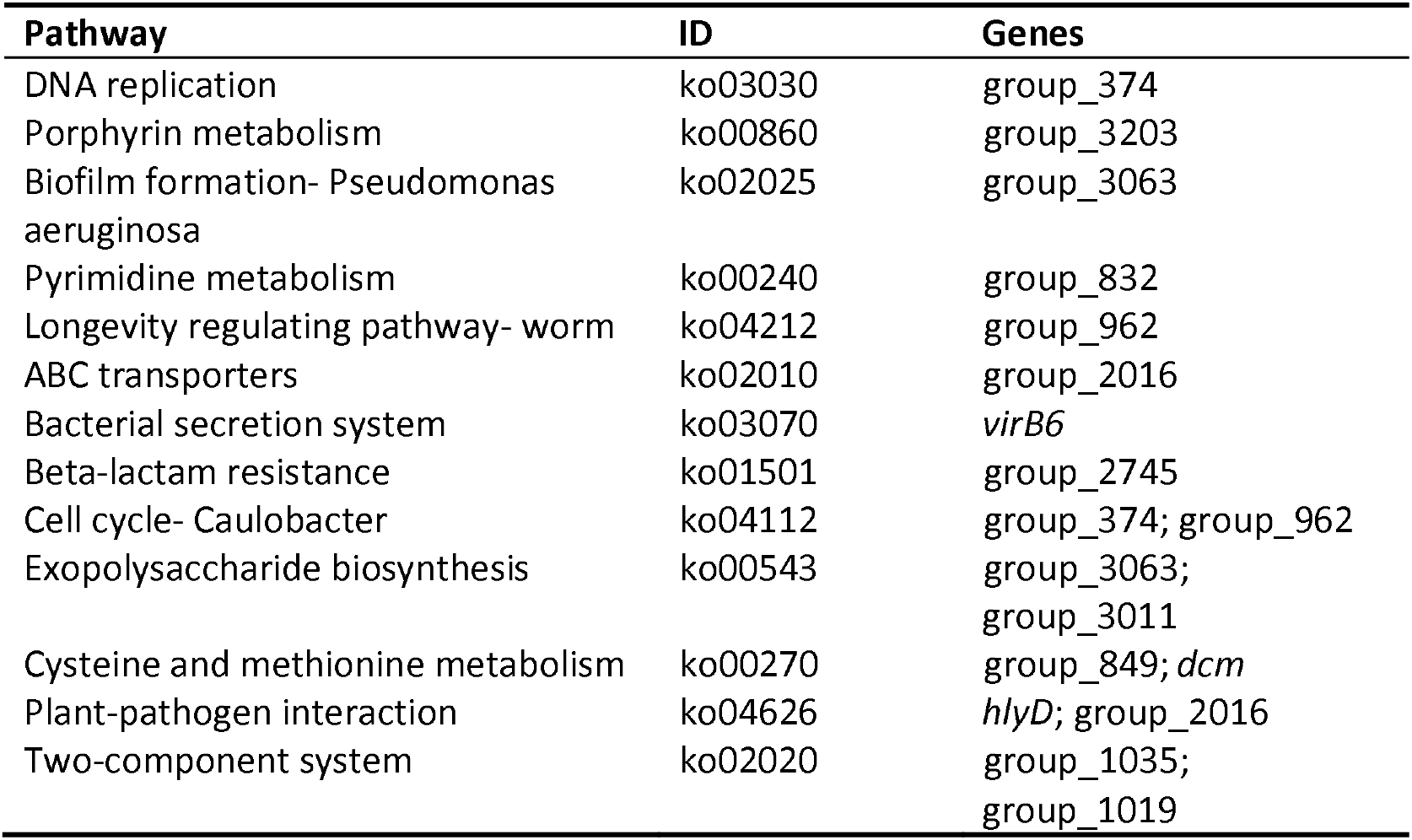
S9-unique genes in the KEGG database and their pathways.

The 53 GO-annotated S9-unqiue genes were enriched according to their gene functions (Fig. 5). The functions associated with more genes are regarded to be more likely to encode higher virulence, but only if these gene functions are related to bacterial virulence. As shown in Fig. 5, the top 10 (ranked by gene numbers) gene functions are cellular metabolic process, primary metabolic process, nitrogen compound metabolic process, organic substance metabolic process, small molecule binding, hydrolase activity, ion binding, transferase activity, heterocyclic compound binding and organic cyclic compound binding. Among these, hydrolase activity and ion binding are most likely suspect because some hydrolases such as the esterase are virulence factors in some bacteria [6-8] and the ion binding function may be associated with efflux pump, involved in drug resistance in bacteria [9,10].

**Figure 5.**
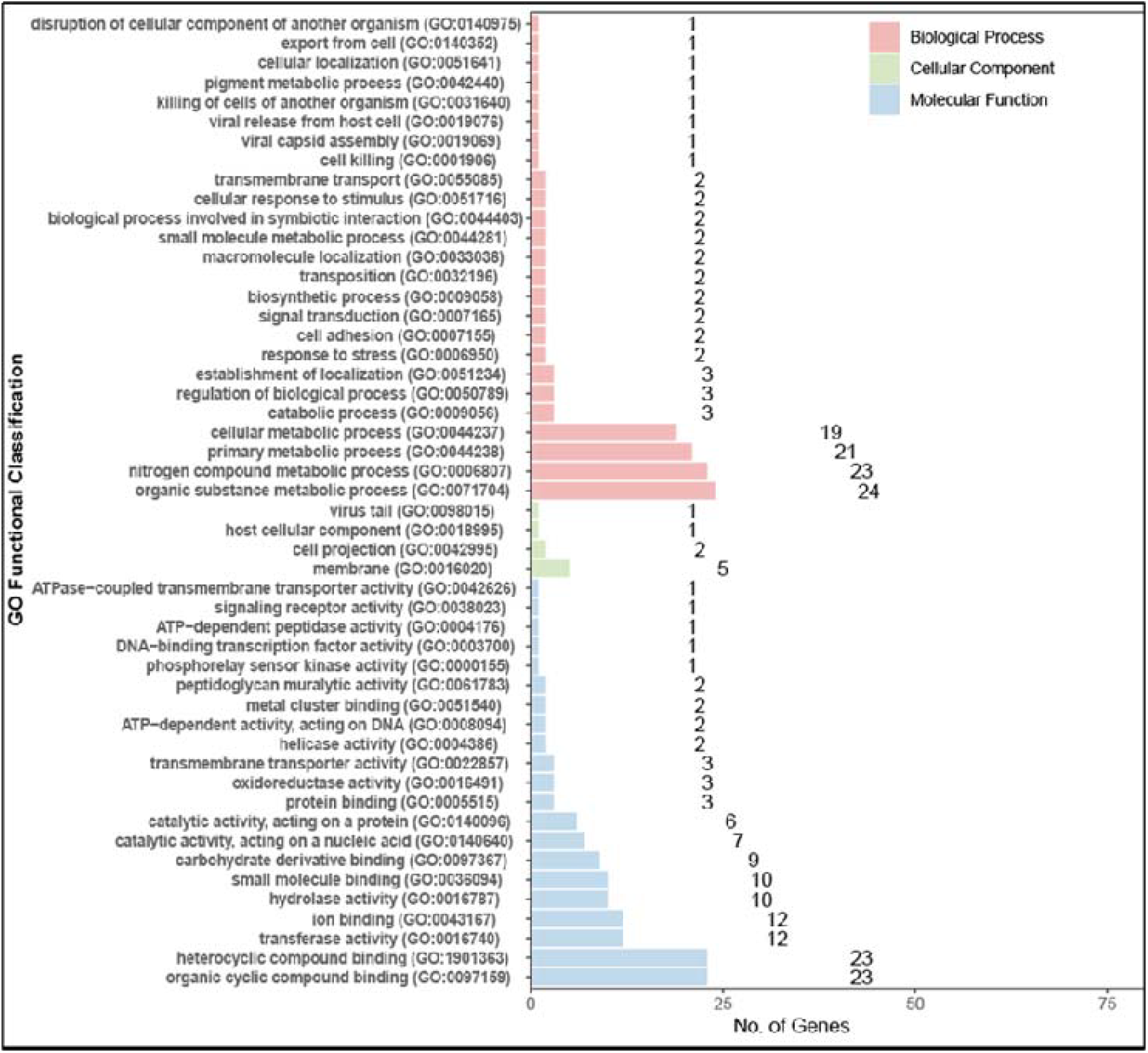
GO functional enrichment of S9-unique genes. In the figure, the items are group by three level 1 GO items: Molecular Function (MF, blue), Cell Component (CC, green) and Biological Process (BP, red).

KEGG enrichment analysis of 14 KEGG-annotated S9-unqiue genes and their associated pathways are listed in Table 2. Several pathways including biofilm formation [11,12], ABC transporters [13], bacterial secretion system [14] and two-component system [15,16] need special attention because they are associated either with drug resistance or with the bacterial virulence. In addition, the genes *virB6, dcm* and *hlyD* also need special attention because *virB*6 is associated with sporty pili and type IV secretion system (virulence factors) of *Agrobacterium* [17], *dcm* is a DNA cytosine methylase gene that may affect the virulence and drug resistance of bacteria [18], and *hlyD* participates in secretion of haemolysin (a virulence factor) in *Escherichia coli* [19,20]. The comparison between GO enrichment and KEGG enrichment shows that the ion binding function and the ABC transporter pathway share the same gene group_2016, and therefore the ten genes associated with the ion binding function should be the candidate genes for future analysis.

### Host response to S. muris infection by transcriptome analysis

THP-1 cells infected by S9 and S8 were subjected to RNA-seq analysis, and their differentially expressed genes (S9-vs-S8, i.e., genes expressed in S9 but not in S8) are shown in Fig. 6. It was found that 40 genes were obviously up-regulated and 13 down-regulated in S9-infected host cells. The GO enrichment analysis of these differentially expressed genes is shown in Fig. 7.

**Figure 6.**
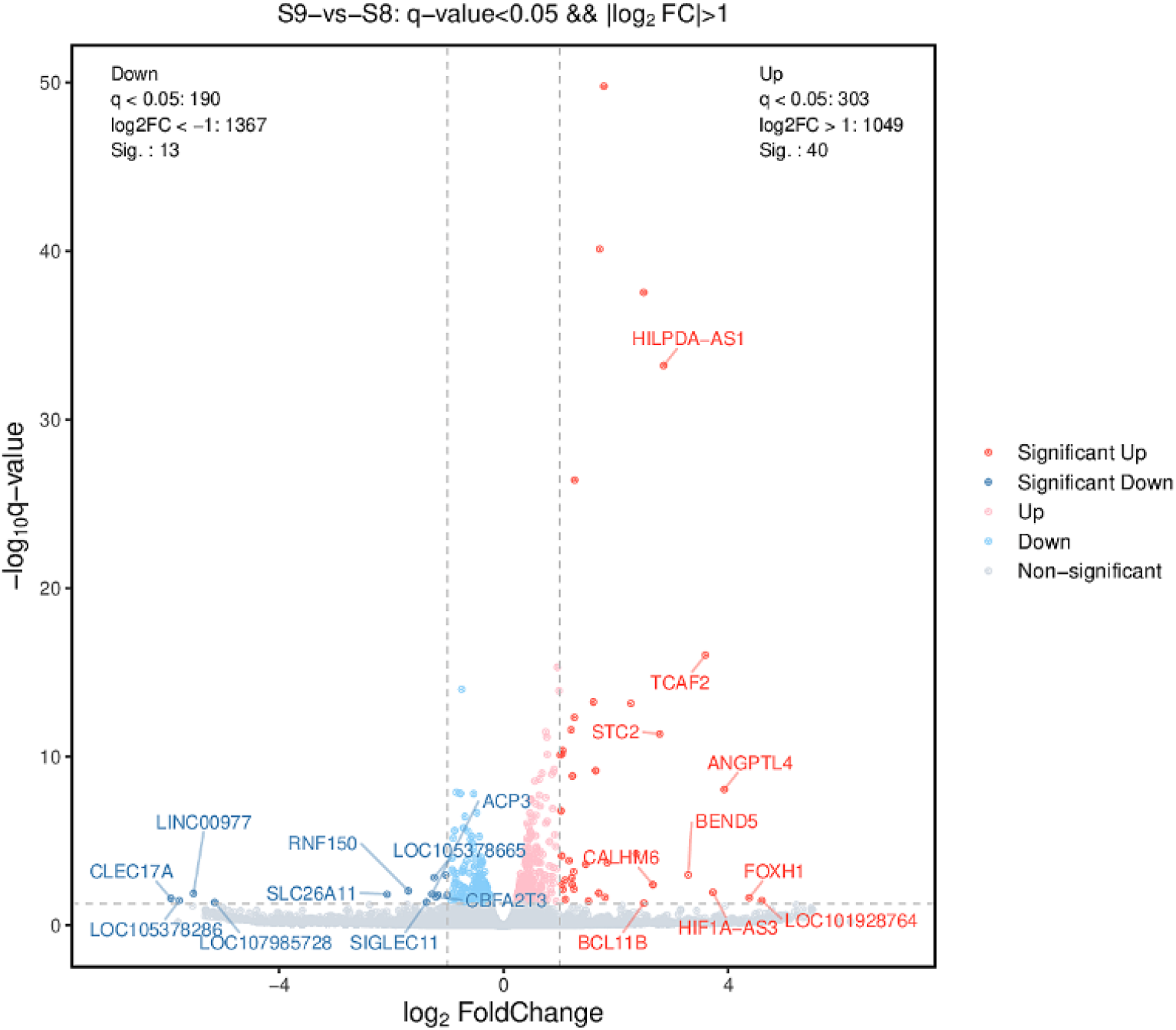
Volcano map of differentially expressed genes (S9-vs-S8). The abscissa is the logarithm (2-based) of the difference multiple (Fold Change, FC), and the ordinate is the negative logarithm of the q-value. Gray points are differentially expressed genes below threshold. Dark blue (points within the interval log_2_FC<-1 and -log_10_q-value>1) and dark red (points within the interval log_2_FC>1 and -log_10_q-value>1) are significantly down-regulated and up-regulated genes, respectively.

**Figure 7.**
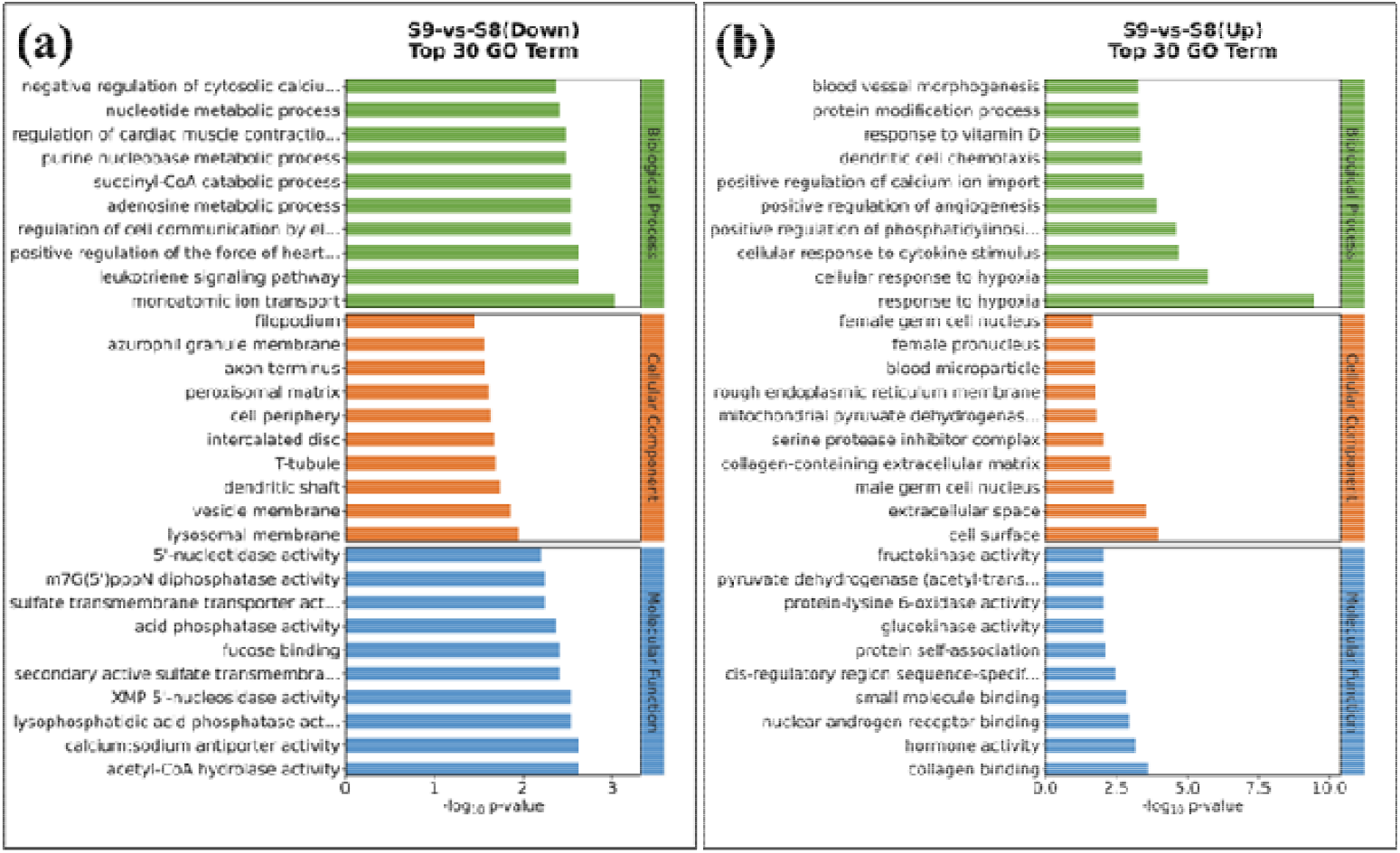
GO enrichment of S9-vs-S8 differentially expressed genes. (a) is for down-regulated genes, and (b) is for up-regulated. Larger log_10_p-value indicates higher enrichment level.

The top 10 (ranked by -log_10_p-value) down-regulated GO terms (gene functions) are monoatomic ion transport, leukotriene signaling pathway, positive regulation of the heart contraction, acetyl-CoA hydrolase activity, calcium: sodium antiporter activity, regulation of cell communication by electrical coupling, adenosine metabolic process, succinyl-CoA catabolic process, lysophosphatidic acid phosphatase activity and XMP 5’-nucleosidase activity and their corresponding genes are *SLC26A11, SLC8A1, CYSLTR1, NUDT7, ACP3*. As discussed above, the ion binding gene function should draw special attention because it may be associated with higher virulence of S9. Here it is found that monoatomic ion transport in S9-infected THP-1 cells is most seriously affected, which echoes the WGS result that the ion binding gene function in S9 could be responsible for higher virulence. The two associated genes of monoatomic ion transport are *SLC26A11* and *SLC8A1*, where *SLC26A11* is related to the transport of chloride ion [21,22] and *SLC8A1* is related to the transport of calcium and sodium ions [23]. The top 2 up-regulated GO terms are response to hypoxia and cellular response to hypoxia. And *SLC26A11* could be the major chloride entry pathway under hypoxia [22]. Thus, it can be inferred that the infection of S9 may hinder calcium and sodium and the oxygen intake of THP-1 cells. The KEGG enrichment analysis also supports the above findings (see Fig. 8). In the down-regulated genes, the calcium signaling pathway has the highest enrichment level, a finding that is consistent with one of the associated gene being *SLC8A1*. In the up-regulated genes, the HIF-1 signaling pathway is at the highest enrichment level. HIF-1, the hypoxia-inducible factor 1, is a helix-loop-helix transcription factor which can activate genes involved in hypoxic homeostasis response proteins [23], which again indicates that infection of S9 may affect the oxygen intake of THP-1 cells.

**Figure 8.**
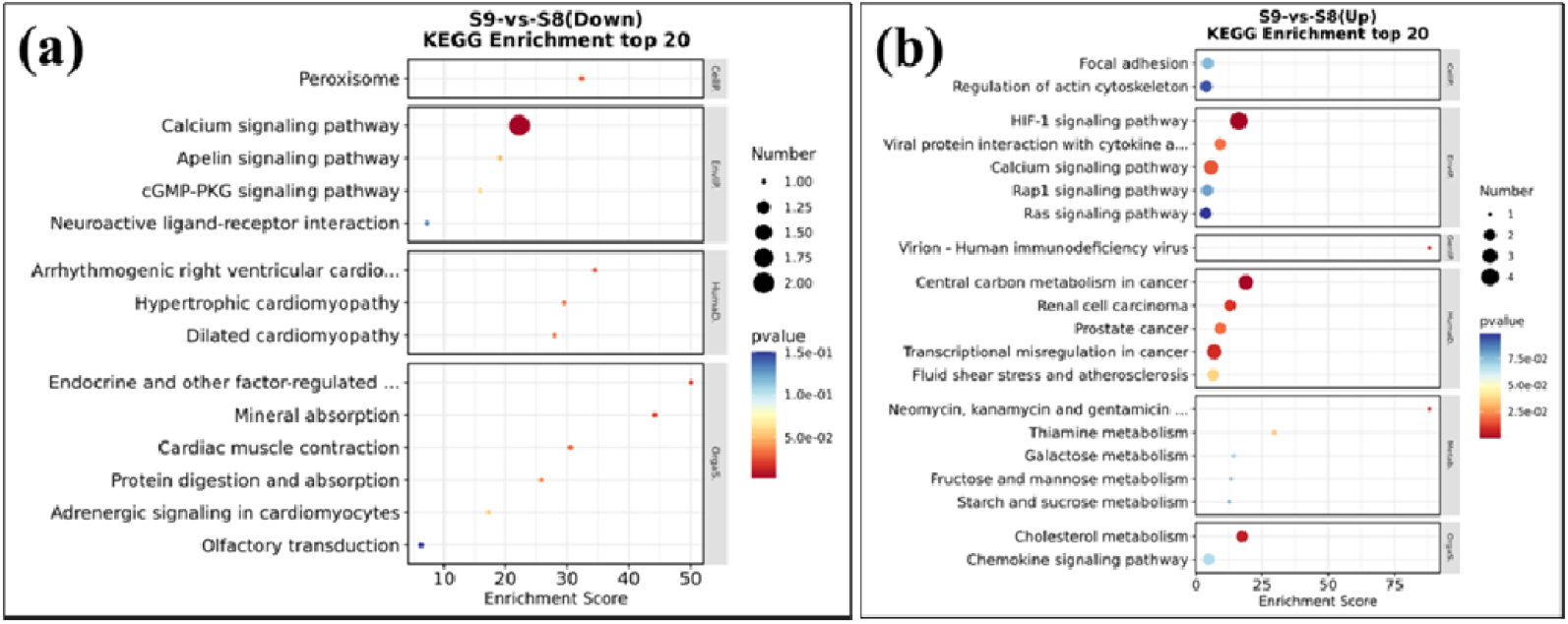
KEGG enrichment of S9-vs-S8 differentially expressed genes. (a) is for down-regulated genes, and (b) is for up-regulated genes. Smaller p-values and larger enrichment scores indicate higher enrichment level.

### S. muris strains are highly resistant to last resort antibiotics colistin

To determine potential treatment response of the *S. muris* strains, we also determined their antibiotic susceptibility to six commonly used antibiotics including ceftazidime, levofloxacin, colistin, polymyxin B, sulfamethoxazole/trimethoprim, minocyclineand ceftazidime for treatment of *S. maltophilia* infections [24]. The results of the antibiotic susceptibility testing for S1, S8 and S9 are listed in Table 3. Except for sulfamethoxazole/trimethoprim (the first-line drug for therapy of *S. maltophilia* infections) and minocycline, the MICs of all the remaining drugs against *S. muris* were much higher than that against *S. maltophilia*, which implies that S. *muris* strains exhibited higher drug resistances and that the treatment of *S. muris* may be more challenging and should be distinguished in the clinic. However, it is of interest to note that both S8 and S9 were quite sensitive to minocycline (MIC<0.06 μg/ml), and minocycline may be a specific and useful medication for the treatment of *S. muris* infections. Further studies are needed to confirm these findings in the clinic.

**Table 3.**
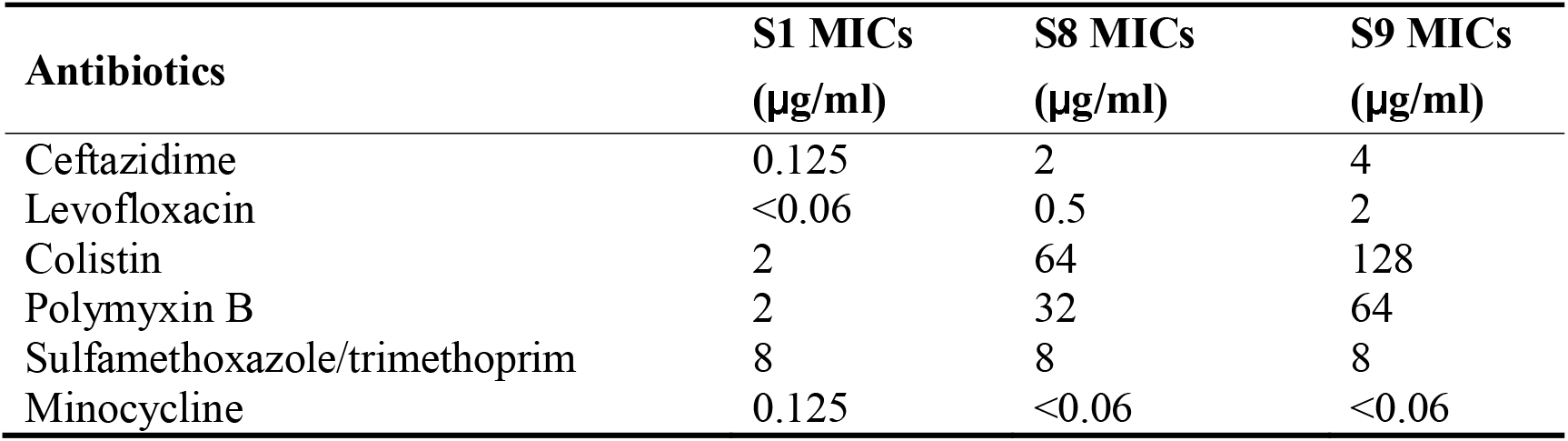
Results of antibiotic susceptibility testing for strains S1, S8 and S9.

## Conclusion

In conclusion, *S. muris* clinical isolates were initially misidentified as *S. maltophilia* by routine mass spectrometry but were subsequently correctly identified as *S. muris by* WGS. The *S. muris s*train S9 isolated from the patient’s blood has stronger virulence than the pulmonary isolate S8 and *S. maltophilia* type strain, which are assocaited with unique candidate genes that may encode the higher virulence including *virB6, dcm, hlyD*, and 14 other genes of unknown function. RNA-seq analysis suggests that the highly virulent *S. muris* strain S9 may preferentially hinder the oxygen intake ion transport and calcium signalling of THP-1 cells. Antibiotic susceptibility testing indicated that compared with *S. maltophilia*, the *S. muris* strains, though more susceptible to minocycline, are highly resistant to last resort antibitoics colistin and polymyxin B and are also resistant to cephalosporin and fluroquinolone antibitoics, which may need to take into consideration when treating *S. muris* infections. Because of the above differences in virulence properties and antibiotic susceptibility, it is critical that *S. muris* be distinguished from *S. maltophilia* for clinical surveillance and for improved treatment outcomes in the future.

## Materials and methods

### Source of bacterial strains and clinical isolates

S1 was *Stenotrophomonas maltophilia* type strain (ATCC13637) from ATCC. S8 was isolated from the sputum of a male patient in his 60s, diagnosed with acute lymphoblastic leukaemia accompanied by pulmonary infection. The antibiotic treatment was not effective and the patient had poor outcome. S9 was isolated from the blood of a male patient in his 20s, diagnosed with septic shock, severe pneumonia, disseminated mucormycosis, and multiple organ failure. S8 and S9 were initially identified as *Stenotrophomonas maltophilia* by standard routine MALDI-TOF mass spectrometry.

### SYTOX Green staining

THP-1 cells were seeded into a 12-well tissue culture plate at the density of 5×10^5^ cells per well. The 12 wells were divided into four groups with 3 wells in each group and Group 1 was the control group with THP-1 cells infected by no bacteria. In Groups 2-4, the cells were infected by S1, S8 and S9 with the multiplicity of infection (MOI=1), respectively. All cells were incubated at 37°C for 18 (or 8) hours and then washed by Hanks’ buffered salt solution (HBSS). Then the washed cells were stained with SYTOX Green (ThermoFisher Scientific) for 30 minutes. Several images were taken by fluorescence microscope with 10× magnification for each well and the images with the best quality were chosen.

### LDH release assay

THP-1 cells were seeded into a 96-well plate and each group had 3 replicates. The control groups (bacterial culture or cells only) were not infected by any bacteria and the three experimental groups were infected by S1, S8 and S9 (MOI=1:1), respectively. After 24-hour incubation at 37°C, the reagent used to lyse cells was added to one of the control groups, and then incubation was continued. After 24 hours, incubation of all groups was stopped. Then THP-1 cells were centrifuged for 5 minutes and the supernatants were transferred to a new 96-well plate. Then, the LDH solution was added into each well and incubated at 37°C for 30 minutes in the dark. Absorbance at 490nm was measured for each well in a Biotek plate reader. The formula for the mortality calculation was described in the caption of Fig. 2. The LDH assay was performed according to the instruction of the LDH kit (Beyotime Biotechnology, Shanghai, China).

### Survival curves of Galleria mellonella

Five groups (20 in each group) of *G. mellonella* larvae with similar health conditions were used. Two groups were the control groups without any bacterial infections, and the other three groups were infected by S1, S8 and S9, respectively. Dead or alive states were recorded daily until the scheduled time. The experiments were stopped after 72 hours. Bacteria were prepared as follows. A single colony was inoculated into Luria-Bertani (LB) broth and incubated at 37°C for 20 hours with shaking. The overnight culture was centrifuged and the pelleted bacteria were resuspended in phosphate-buffered saline (PBS) at a concentration of 1×10^8^ CFU/mL. *G. mellonella* larvae were infected by injection of 10 μL of the bacterial suspension (with the final inoculum size 1×10^7^ CFU per *G. mellonella* larva). One control group was injected with nothing and the other control group was injected with 10 μL of PBS.

### Survival curves of infected mice

Female Balb/c mice aged 6∼8 weeks were used in this experiment. Bacterial inocula were prepared with similar procedures as that used in the *G. mellonella* larvae test. Three groups (10 in each group) of mice were infected via nasal instillation by 50 μL of suspensions (final inoculum size of 1×10^8^ CFU per mouse) of S1, S8 and S9, respectively. All mice were fed in the same environment with *ad libitum* access to food and water. Dead or alive states of infected mice were recorded daily for 7 days.

### Whole genome sequencing (WGS)

AxyPrep bacterial genomic DNA miniprep kit (Axygen Scientific, Union City, CA, USA) was used to extract DNA of the target bacteria. The Illumina HiSeq 2500 platform (paired-end run; 2 × 150 bp) together with an Oxford Nanopore MinION platform was employed to perform the whole-genome sequencing. The Assembly and SRA databases of NCBI were utilized to obtain the publicly available genome assemblies and short-read data for S1, S8 and S9. Kingfisher v7.6.1 (https://github.com/onevcat/Kingfisher) and ncbi-genome-download v0.3.1 (https://github.com/kblin/ncbi-genome-download) were used during the data retrieval. Fastp v0.23.2 was used to filter qualities and trim adaptors of sequencing reads. Careful-mode shovill v1.1.0 (https://github.com/tseemann/shovill) was used to assemble trimmed reads and contigs which possess less than 200 bp. With the help of Unicycler hybrid assembly pipeline, long-read data from Oxford Nanopore MinION sequencing and short-read data from Illumina sequencing were synergized to generate the complete genomes of S1, S8 and S9. ANI was utilized to identify species of S8 and S9.

### RNA-seq analysis of infected host cells

Six groups of THP-1 cells were prepared. Three groups (S8-1, S8-2 and S8-3) were infected by S8 and three groups (S9-1, S9-2 and S9-3) were infected by S9 (the MOIs were all 1:1 and the infection time was 24 hours). The total RNAs of these six groups of THP-1 cells (after infections) were extracted using total RNA Trizol kit (Beyotime Biotechnology, Shanghai, China). Before sequencing, rRNAs were removed from the total RNAs, then the remaining RNAs were converted into cDNAs by reverse transcription. The sequencing method of cDNA was the same as DNA in WGS.

### Antibiotic susceptibility testing

The standard microdilution method was used to determine the minimum inhibitory concentration (MIC). *S. maltophilia* ATCC13637 (S1), and *S. muris* S8, S9 were cultured in LB broth with shaking overnight at 37°C. The bacterial suspension was adjusted to 0.5 McFarland and then diluted 1:100 in CAMHB medium. Then 100 μL diluted culture was transferred to 96-well microtiter plates and mixed with serial two-fold dilutions of ceftazidime, levofloxacin, colistinsulfate, polymyxin B, minocycline and sulfamethoxazole/ trimethoprim in concentrations ranging from 128 μg/mL to 0.06 μg/mL. The negative control contained only CAMHB, and the positive control contained diluted culture in CAMHB. Assay plates were incubated without shaking at 37□°C for 20□h. The MIC was the lowest concentration of each drug where no visible growth was seen in the wells. All experiments were run in triplicate.

## Data Availability

All data produced in the present study are available upon reasonable request to the authors

## Acknowledgement

We thank Novogene (Beijing, China, https://cn.novogene.com/) and OE Biotech (https://www.oebiotech.com/) for the sequencing service.

## Author contributions

Y. Z and J.Y. L designed and conceived this work. J. Y. L and Y. H. X performed experiments. J. Y. L, X. D, Y. Y. Y, T. T. W, Y. H. X, X. Y analysed data. J. Y. L and Y. Z wrote and finalized the manuscript. All authors read, revised and approved the manuscript.

## References

[1] Diard M, Hardt WD. Evolution of bacterial virulence. FEMS Microbiol Rev 2017; 41(5):679–97. 10.1093/femsre/fux023

[2] Pompilio A, Crocetta V, De Nicola S, et al. Cooperative pathogenicity in cystic fibrosis: Stenotrophomonas maltophilia modulates Pseudomonas aeruginosa virulence in mixed biofilm. Front Microbiol 2015; 6(951. 10.3389/fmicb.2015.00951

[3] Wicaksono WA, Erschen S, Krause R, et al. Enhanced survival of multi-species biofilms under stress is promoted by low-abundant but antimicrobial-resistant keystone species. J Hazard Mater 2022; 422(126836. 10.1016/j.jhazmat.2021.126836

[4] Zhao J, Grant SF. Advances in whole genome sequencing technology. Curr Pharm Biotechnol 2011; 12(2):293–305. 10.2174/138920111794295729

[5] Afrizal A, Jennings SAV, Hitch TCA, et al. Enhanced cultured diversity of the mouse gut microbiota enables custom-made synthetic communities. Cell Host Microbe 2022; 30(11):1630–45 e25. 10.1016/j.chom.2022.09.011

[6] Songer JG. Bacterial phospholipases and their role in virulence. Trends Microbiol 1997; 5(4):156–61. 10.1016/S0966-842X(97)01005-6

[7] Thomas R, Hamat RA, Neela V. Extracellular enzyme profiling of Stenotrophomonas maltophilia clinical isolates. Virulence 2014; 5(2):326–30. 10.4161/viru.27724

[8] Travassos LH, Pinheiro MN, Coelho FS, et al. Phenotypic properties, drug susceptibility and genetic relatedness of Stenotrophomonas maltophilia clinical strains from seven hospitals in Rio de Janeiro, Brazil. J Appl Microbiol 2004; 96(5):1143–50. 10.1111/j.1365-2672.2004.02248.x

[9] Hu RM, Liao ST, Huang CC, et al. An inducible fusaric acid tripartite efflux pump contributes to the fusaric acid resistance in Stenotrophomonas maltophilia. PLoS One 2012; 7(12):e51053. 10.1371/journal.pone.0051053

[10] Huang YW, Hu RM, Chu FY, et al. Characterization of a major facilitator superfamily (MFS) tripartite efflux pump EmrCABsm from Stenotrophomonas maltophilia. J Antimicrob Chemother 2013; 68(11):2498–505. 10.1093/jac/dkt250

[11] Jefferson KK. What drives bacteria to produce a biofilm? FEMS Microbiol Lett 2004; 236(2):163–73. 10.1016/j.femsle.2004.06.005

[12] Olsen I. Biofilm-specific antibiotic tolerance and resistance. Eur J Clin Microbiol Infect Dis 2015; 34(5):877–86. 10.1007/s10096-015-2323-z

[13] Zajac OM, Tyski S, Laudy AE. The Contribution of Efflux Systems to Levofloxacin Resistance in Stenotrophomonas maltophilia Clinical Strains Isolated in Warsaw, Poland. Biology (Basel) 2022; 11(7) 10.3390/biology11071044

[14] Nas MY, White RC, DuMont AL, et al. Stenotrophomonas maltophilia Encodes a VirB/VirD4 Type IV Secretion System That Modulates Apoptosis in Human Cells and Promotes Competition against Heterologous Bacteria, Including Pseudomonas aeruginosa. Infect Immun 2019; 87(9) 10.1128/IAI.00457-19

[15] Gooderham WJ, Hancock RE. Regulation of virulence and antibiotic resistance by two-component regulatory systems in Pseudomonas aeruginosa. FEMS Microbiol Rev 2009; 33(2):279–94. 10.1111/j.1574-6976.2008.00135.x

[16] Lei L, Long L, Yang X, et al. The VicRK Two-Component System Regulates Streptococcus mutans Virulence. Curr Issues Mol Biol 2019; 32(167-200. 10.21775/cimb.032.167

[17] Judd PK, Mahli D, Das A. Molecular characterization of the Agrobacterium tumefaciens DNA transfer protein VirB6. Microbiology (Reading) 2005; 151(Pt 11):3483–92. 10.1099/mic.0.28337-0

[18] Gao Q, Lu S, Wang Y, et al. Bacterial DNA methyltransferase: A key to the epigenetic world with lessons learned from proteobacteria. Front Microbiol 2023; 14(1129437. 10.3389/fmicb.2023.1129437

[19] Moeinizadeh H, Shaheli M. Frequency of hlyA, hlyB, hlyC and hlyD genes in uropathogenic Escherichia coli isolated from UTI patients in Shiraz. GMS Hyg Infect Control 2021; 16(Doc25. 10.3205/dgkh000396

[20] Schulein R, Gentschev I, Schlor S, et al. Identification and characterization of two functional domains of the hemolysin translocator protein HlyD. Mol Gen Genet 1994; 245(2):203–11. 10.1007/BF00283268

[21] Rahmati N, Vinueza Veloz MF, Xu J, et al. SLC26A11 (KBAT) in Purkinje Cells Is Critical for Inhibitory Transmission and Contributes to Locomotor Coordination. eNeuro 2016; 3(3) 10.1523/ENEURO.0028-16.2016

[22] Wei S, Chen B, Low SW, et al. SLC26A11 Inhibition Reduces Oncotic Neuronal Death and Attenuates Stroke Reperfusion Injury. Mol Neurobiol 2023; 60(10):5931–43. 10.1007/s12035-023-03453-1

[23] Liu K, Liu Z, Qi H, et al. Genetic Variation in SLC8A1 Gene Involved in Blood Pressure Responses to Acute Salt Loading. Am J Hypertens 2018; 31(4):415–21. 10.1093/ajh/hpx179

[24] Liu J, Xiang Y, Zhang Y. Stenotrophomonas maltophilia: An Urgent Threat with Increasing Antibiotic Resistance. Current Microbiology 2023; 81(1):6. 10.1007/s00284-023-03524-5

