## Supplementary Table S1 and Table S2 for "*Stenotrophomonas muris* - First discovered as an urgent human pathogen with strong virulence associated with bloodstream infections": Supplementary material-Tables.docx

**Table S1** Genes of S1, S8 and S9 in VFDB database (with identity > 60%)

| S1 genes | Identity (%) | S8 genes | Identity (%) | S9 genes | Identity (%) |
| --- | --- | --- | --- | --- | --- |
| PA3349 | 65.55 | htpB | 64.62 | nueA | 67.82 |
| flgG | 69.12 | acrB | 68.98 | katA | 76.22 |
| flgI | 71.49 | waaA | 67 | pilU | 77.62 |
| fliG | 69.09 | flmH | 69.96 | pilT | 80.37 |
| fliM | 74.83 | acpXL | 74.07 | pilZ | 72.37 |
| fliP | 71.84 | pilZ | 71.77 | acpXL | 74.07 |
| flhA | 72.02 | tapT | 74.82 | flmH | 69.65 |
| flhG | 70.85 | pilU | 77.81 | cheW | 75.94 |
| cheY | 76.69 | katA | 76.37 | motC | 70.58 |
| motC | 69.31 | nueA | 67.67 | cheY | 77.81 |
| cheW | 75.06 | kdsA | 65.55 | flhG | 71 |
| cheB | 66.7 | motA | 67.95 | flhA | 71.89 |
| xcpA/pilD | 68.06 | pilU | 71.35 | fliP | 73.16 |
| tapC | 65.82 | algA | 69.31 | fliM | 74.46 |
| pilR | 71.06 | rfbK1 | 66.19 | fliG | 69.73 |
| pilG | 79.53 | flgG | 69.79 | flgI | 71.08 |
| flmH | 69.65 | flgI | 70.9 | flgG | 69.92 |
| acpXL | 74.07 | icl | 73.76 | pilM | 68.31 |
| pilZ | 72.67 | algW | 65.04 | motA | 67.95 |
| pilT | 79.3 | pilR | 72.2 | pilU | 71.61 |
| pilU | 77.13 | tapC | 65.66 | algA | 69.38 |
| katA | 75.84 | xcpA/pilD | 68.64 | rfbK1 | 66.19 |
| acrB | 68.87 | pilM | 68.61 | cheB | 67.8 |
| htpB | 64.71 | cheB | 67.63 | cheR | 68.09 |
| rfbK1 | 68.43 | cheR | 67.7 | acrB | 69.11 |
| algA | 68.55 | xcpT | 69.92 | htpB | 64.69 |
| pilU | 71.65 | pilG | 80.42 | icl | 74.25 |
| narH | 71.42 | pilH | 69.38 | algW | 65.04 |
| xcpT | 70.49 | fliG | 69.68 | pilH | 70 |
| tufA | 67.61 | fliM | 74.46 | waaA | 67 |
| tufA | 67.73 | fliP | 72.83 | pilR | 71.84 |
| pilM | 69.67 | flhA | 71.67 | xcpA/pilD | 68.77 |
| icl | 74.61 | flhG | 71.17 | pilG | 80.42 |
| pchD | 68.41 | cheY | 77.53 |  |  |
| waaA | 66.19 | motC | 70.74 |  |  |
|  |  | cheW | 75.69 |  |  |

**TABLE S2** S9-unique genes (the S1-repeated genes were omitted) in GO database

| Gene_ID | GO_term | Function_class | Function |
| --- | --- | --- | --- |
| dcm_orf5049 | GO:0003824 | molecular_function | catalytic activity |
| group_1014_orf4779 | GO:0003824 | molecular_function | catalytic activity |
| group_1019_orf4993 | GO:0009987 | biological_process | cellular process |
| group_1019_orf4993 | GO:0110165 | cellular_component | cellular anatomical entity |
| group_1035_orf4930 | GO:0009987 | biological_process | cellular process |
| group_1035_orf4930 | GO:0110165 | cellular_component | cellular anatomical entity |
| group_1077_orf5011 | GO:0003824 | molecular_function | catalytic activity |
| group_1165_orf1487 | GO:0003824 | molecular_function | catalytic activity |
| group_1165_orf1487 | GO:0005488 | molecular_function | binding |
| group_1165_orf1487 | GO:0008152 | biological_process | metabolic process |
| group_1165_orf1487 | GO:0009987 | biological_process | cellular process |
| group_1239_orf1295 | GO:0003824 | molecular_function | catalytic activity |
| group_1239_orf1295 | GO:0005488 | molecular_function | binding |
| group_1239_orf1295 | GO:0008152 | biological_process | metabolic process |
| group_1239_orf1295 | GO:0009987 | biological_process | cellular process |
| group_1285_orf647 | GO:0003824 | molecular_function | catalytic activity |
| group_1285_orf647 | GO:0005488 | molecular_function | binding |
| group_1286_orf1369 | GO:0003824 | molecular_function | catalytic activity |
| group_1286_orf1369 | GO:0005488 | molecular_function | binding |
| group_1286_orf1369 | GO:0008152 | biological_process | metabolic process |
| group_1286_orf1369 | GO:0009987 | biological_process | cellular process |
| group_1286_orf1369 | GO:0140657 | molecular_function | ATP-dependent activity |
| group_1317_orf1396 | GO:0003824 | molecular_function | catalytic activity |
| group_1317_orf1396 | GO:0005488 | molecular_function | binding |
| group_1317_orf1396 | GO:0008152 | biological_process | metabolic process |
| group_1317_orf1397 | GO:0005488 | molecular_function | binding |
| group_1345_orf1100 | GO:0016032 | biological_process | viral process |
| group_1352_orf1175 | GO:0003824 | molecular_function | catalytic activity |
| group_1352_orf1175 | GO:0005488 | molecular_function | binding |
| group_1352_orf1175 | GO:0008152 | biological_process | metabolic process |
| group_1352_orf1175 | GO:0009987 | biological_process | cellular process |
| group_1383_orf1410 | GO:0005488 | molecular_function | binding |
| group_1383_orf1410 | GO:0008152 | biological_process | metabolic process |
| group_1383_orf1410 | GO:0009987 | biological_process | cellular process |
| group_1383_orf1410 | GO:0050896 | biological_process | response to stimulus |
| group_1462_orf536 | GO:0065007 | biological_process | biological regulation |
| group_1462_orf536 | GO:0140110 | molecular_function | transcription regulator activity |
| group_1757_orf1785 | GO:0003824 | molecular_function | catalytic activity |
| group_1757_orf1785 | GO:0008152 | biological_process | metabolic process |
| group_1757_orf1785 | GO:0009987 | biological_process | cellular process |
| group_1757_orf1785 | GO:0065007 | biological_process | biological regulation |
| group_1757_orf1785 | GO:0140299 | molecular_function | small molecule sensor activity |
| group_1966_orf4519 | GO:0005488 | molecular_function | binding |
| group_1966_orf4519 | GO:0008152 | biological_process | metabolic process |
| group_1966_orf4519 | GO:0009987 | biological_process | cellular process |
| group_1966_orf4520 | GO:0005488 | molecular_function | binding |
| group_1966_orf4520 | GO:0008152 | biological_process | metabolic process |
| group_1966_orf4520 | GO:0009987 | biological_process | cellular process |
| group_2016_orf3846 | GO:0003824 | molecular_function | catalytic activity |
| group_2016_orf3846 | GO:0005215 | molecular_function | transporter activity |
| group_2016_orf3846 | GO:0005488 | molecular_function | binding |
| group_2016_orf3846 | GO:0008152 | biological_process | metabolic process |
| group_2016_orf3846 | GO:0009987 | biological_process | cellular process |
| group_2016_orf3846 | GO:0051179 | biological_process | localization |
| group_2016_orf3846 | GO:0110165 | cellular_component | cellular anatomical entity |
| group_2016_orf3846 | GO:0140657 | molecular_function | ATP-dependent activity |
| group_2148_orf3752 | GO:0008152 | biological_process | metabolic process |
| group_2259_orf3945 | GO:0003824 | molecular_function | catalytic activity |
| group_2259_orf3945 | GO:0005488 | molecular_function | binding |
| group_2355_orf4489 | GO:0005488 | molecular_function | binding |
| group_2361_orf4077 | GO:0005488 | molecular_function | binding |
| group_2361_orf4077 | GO:0008152 | biological_process | metabolic process |
| group_2361_orf4077 | GO:0009987 | biological_process | cellular process |
| group_2368_orf4088 | GO:0003824 | molecular_function | catalytic activity |
| group_2368_orf4088 | GO:0005488 | molecular_function | binding |
| group_2368_orf4088 | GO:0008152 | biological_process | metabolic process |
| group_2368_orf4088 | GO:0009987 | biological_process | cellular process |
| group_2745_orf4196 | GO:0005215 | molecular_function | transporter activity |
| group_2745_orf4196 | GO:0009987 | biological_process | cellular process |
| group_2745_orf4196 | GO:0051179 | biological_process | localization |
| group_2745_orf4196 | GO:0110165 | cellular_component | cellular anatomical entity |
| group_2810_orf4237 | GO:0003824 | molecular_function | catalytic activity |
| group_2810_orf4237 | GO:0005488 | molecular_function | binding |
| group_2810_orf4237 | GO:0008152 | biological_process | metabolic process |
| group_2810_orf4237 | GO:0009987 | biological_process | cellular process |
| group_2942_orf2107 | GO:0005215 | molecular_function | transporter activity |
| group_2942_orf2107 | GO:0110165 | cellular_component | cellular anatomical entity |
| group_3011_orf2123 | GO:0003824 | molecular_function | catalytic activity |
| group_3203_orf1996 | GO:0003824 | molecular_function | catalytic activity |
| group_3203_orf1996 | GO:0008152 | biological_process | metabolic process |
| group_3203_orf1996 | GO:0009987 | biological_process | cellular process |
| group_342_orf1584 | GO:0008152 | biological_process | metabolic process |
| group_342_orf1584 | GO:0009987 | biological_process | cellular process |
| group_342_orf1584 | GO:0065007 | biological_process | biological regulation |
| group_347_orf1673 | GO:0003824 | molecular_function | catalytic activity |
| group_347_orf1673 | GO:0005488 | molecular_function | binding |
| group_374_orf104 | GO:0003824 | molecular_function | catalytic activity |
| group_374_orf104 | GO:0005488 | molecular_function | binding |
| group_374_orf104 | GO:0008152 | biological_process | metabolic process |
| group_374_orf104 | GO:0009987 | biological_process | cellular process |
| group_374_orf104 | GO:0140657 | molecular_function | ATP-dependent activity |
| group_603_orf15 | GO:0005488 | molecular_function | binding |
| group_603_orf15 | GO:0008152 | biological_process | metabolic process |
| group_603_orf15 | GO:0009987 | biological_process | cellular process |
| group_634_orf1618 | GO:0003824 | molecular_function | catalytic activity |
| group_634_orf1618 | GO:0005488 | molecular_function | binding |
| group_634_orf1618 | GO:0008152 | biological_process | metabolic process |
| group_634_orf1618 | GO:0009987 | biological_process | cellular process |
| group_832_orf2454 | GO:0003824 | molecular_function | catalytic activity |
| group_832_orf2454 | GO:0008152 | biological_process | metabolic process |
| group_832_orf2454 | GO:0009987 | biological_process | cellular process |
| group_835_orf3533 | GO:0005488 | molecular_function | binding |
| group_835_orf3533 | GO:0060089 | molecular_function | molecular transducer activity |
| group_840_orf2544 | GO:0003824 | molecular_function | catalytic activity |
| group_840_orf2544 | GO:0005488 | molecular_function | binding |
| group_840_orf2544 | GO:0008152 | biological_process | metabolic process |
| group_840_orf2544 | GO:0009987 | biological_process | cellular process |
| group_849_orf2314 | GO:0003824 | molecular_function | catalytic activity |
| group_857_orf3358 | GO:0005488 | molecular_function | binding |
| group_857_orf3358 | GO:0044419 | biological_process | biological process involved in interspecies interaction between organisms |
| group_857_orf3358 | GO:0044423 | cellular_component | virion component |
| group_857_orf3358 | GO:0110165 | cellular_component | cellular anatomical entity |
| group_873_orf3634 | GO:0005488 | molecular_function | binding |
| group_879_orf2616 | GO:0003824 | molecular_function | catalytic activity |
| group_879_orf2616 | GO:0005488 | molecular_function | binding |
| group_911_orf2666 | GO:0003824 | molecular_function | catalytic activity |
| group_911_orf2666 | GO:0005488 | molecular_function | binding |
| group_926_orf2951 | GO:0003824 | molecular_function | catalytic activity |
| group_926_orf2951 | GO:0008152 | biological_process | metabolic process |
| group_926_orf2951 | GO:0009987 | biological_process | cellular process |
| group_926_orf2951 | GO:0016032 | biological_process | viral process |
| group_926_orf2951 | GO:0044419 | biological_process | biological process involved in interspecies interaction between organisms |
| group_943_orf3011 | GO:0003824 | molecular_function | catalytic activity |
| group_943_orf3011 | GO:0005488 | molecular_function | binding |
| group_957_orf3376 | GO:0003824 | molecular_function | catalytic activity |
| group_957_orf3376 | GO:0005488 | molecular_function | binding |
| group_962_orf3117 | GO:0003824 | molecular_function | catalytic activity |
| group_962_orf3117 | GO:0008152 | biological_process | metabolic process |
| group_962_orf3117 | GO:0140657 | molecular_function | ATP-dependent activity |
| group_966_orf2674 | GO:0005488 | molecular_function | binding |
| group_966_orf2674 | GO:0008152 | biological_process | metabolic process |
| group_966_orf2674 | GO:0009987 | biological_process | cellular process |
| group_986_orf2975 | GO:0009987 | biological_process | cellular process |
| group_986_orf2975 | GO:0016032 | biological_process | viral process |
| group_986_orf2975 | GO:0044419 | biological_process | biological process involved in interspecies interaction between organisms |
| higA_orf1544 | GO:0005488 | molecular_function | binding |
| hlyD_orf3802 | GO:0009987 | biological_process | cellular process |
| hlyD_orf3802 | GO:0051179 | biological_process | localization |
| hlyD_orf3802 | GO:0110165 | cellular_component | cellular anatomical entity |
| sRAP_orf4636 | GO:0005488 | molecular_function | binding |
| sRAP_orf4636 | GO:0008152 | biological_process | metabolic process |
| sRAP_orf4636 | GO:0009987 | biological_process | cellular process |
| sRAP_orf4636 | GO:0050896 | biological_process | response to stimulus |
| trbL_orf4064 | GO:0009987 | biological_process | cellular process |
| trbL_orf4064 | GO:0051179 | biological_process | localization |
| virB6_orf4048 | GO:0009987 | biological_process | cellular process |
| virB6_orf4048 | GO:0051179 | biological_process | localization |
